# Bridging mental health, cognition and the brain in mild traumatic brain injury: A multilayer network analysis of the TRACK-TBI study

**DOI:** 10.1101/2025.02.16.25322263

**Authors:** Juan F. Domínguez D., Mervyn Singh, Lyndon Firman-Sadler, Jade Guarnera, Ivan L. Simpson-Kent, Phoebe Imms, Andrei Irimia, Karen Caeyenberghs, TRACK-TBI Investigators

**Affiliations:** Cognitive Neuroscience Unit, School of Psychology, Deakin University, Melbourne, VIC, Australia; Cumming School of Medicine, University of Calgary, Canada; Institute of Psychology, Developmental and Educational Psychology Unit, Leiden University, Leiden, the Netherlands; Department of Psychology, University of Pennsylvania, Philadelphia, PA, USA; Ethel Percy Andrus Gerontology Centre, Leonard Davis School of Gerontology, University of Southern California, Los Angeles, CA, USA; Department of Biomedical Engineering, Viterbi School of Engineering, University of Southern California, Los Angeles, CA, USA; Department of Quantitative & Computational Biology, Dana and David Dornsife College of Arts & Sciences, University of Southern California, Los Angeles, CA, USA

## Abstract

**Background:** Mild traumatic brain injury (mTBI) patients suffer from several mental health symptoms (*e.g*., anxiety, depressive symptoms) and cognitive deficits (*e.g*., attentional deficits, slowed processing speed). However, symptoms in TBI are largely investigated in isolation, using univariate approaches, ignoring the interactions between symptoms and the underlying large-scale brain networks. We constructed the first multilayer network in mTBI to examine the relationships between networks of cognition, mental health and structural brain measures and to identify key variables bridging relationships across these networks.

**Methods:** Chronic phase cross-sectional data (6-month follow-up) from 457 mTBI patients was extracted from the TRACK-TBI Longitudinal study. We selected four variables from self-report mental health questionnaires (affective layer), eight cognitive test scores from the NIH toolbox (cognitive layer), and grey matter volumes from eight brain regions of the central executive and salience networks from anatomical MRI scans (brain layer). We used a multilayer network approach to examine the relationships (edges) between all variables (nodes) across layers. We then used the bridge strength centrality metric to identify nodes that ‘bridge’ the affective, cognitive, and brain layers.

**Results:** Insomnia severity, immediate verbal memory, somatisation and processing speed nodes exceeded an *a priori* 80th percentile threshold on the bridge strength scores and can therefore be regarded as key nodes bridging relationships across affective, cognitive and brain layers.

**Conclusions:** The bridging nodes identified in our multilayer network analyses suggest targets for future studies to develop more customized, efficient, and efficacious treatments to alleviate mental health symptoms and cognitive deficits in mTBI patients.

## INTRODUCTION

In 2019, there were approximately 12.3 million incident cases of mild traumatic injury (mTBI) worldwide.^1^ Between 20-40% of mTBI patients experience long-term deficits.^2^ A number of studies have reported reduced performance in mTBI patients on cognitive tasks assessing attention, memory, attention switching, and executive functioning,^3–7^ together with increased mental health symptoms (depression, anxiety, stress),^8,9^ that present long-term (> 3 months) post injury. Moreover, studies have demonstrated bidirectional relationships between cognitive function and mental health in TBI.^10,11^ These chronic impairments are considered the result of disruptions in the coordinated activity of three neural networks (*i.e.*, the default mode network, salience network, and central executive network.^12–16^

There is mounting evidence from network psychometrics that mental health issues result from the interaction between multiple symptoms in a symptom network (*e.g.*, depressed mood, fatigue, insomnia, difficulty in concentration, motivation), with disruption to a symptom spreading across the network.^17–21^ In the context of moderate-severe TBI, Carmichael and colleagues^18^ showed that the symptoms of worrying and having difficulty relaxing are especially important to the maintenance and comorbidity of post-TBI anxiety and depression. Despite the growing recognition of symptoms as network phenomena, when attempting to ascertain how disruption to the brain networks leads to specific symptoms, studies have focused on isolated behavioural measures that are not in the same statistical framework as the brain measures. It is, however, important to analyse brain-behaviour relationships using the same framework.^22^

Multilayer network analysis is a new approach that allows analysis of brain-behaviour relationships within the same framework: the relationships among variables across levels of organisation, such as behaviour/symptoms and brain variables are modelled simultaneously by placing them all in the same integrated network.^23^ Variables are represented as nodes within multiple layers of this network and their relationships are modelled as correlations between nodes within the same layer and across different layers. Centrality metrics (*e.g.*, bridge strength centrality) are then used to identify nodes that ‘bridge’ the brain and behavioural layers. These bridge nodes exhibit particularly strong relationships with nodes from layers different from their own and thus may serve as previously unrecognized targets for intervention.^24^ Multilayer network analysis is still in its infancy and, to our knowledge, only three other studies have used this approach to study brain-psychometric relationships, in depression,^25^ autism,^26^ and intelligence in struggling learners.^27^

In the present study, we employed, for the first time, this innovative multilayer network approach to examine the relationships between networks of mental health measures, cognitive scores, and grey matter brain volumes (GMV) in chronic mTBI patients, using a subset of the Transforming Research and Clinical Knowledge in TBI Longitudinal (TRACK-TBI LONG) dataset.^28–30^ Our aim was to characterize the complex interactions between behavioural symptoms and underlying brain mechanisms in mTBI patients. More specifically, we first wanted to apply a network psychometrics approach to model mental health (*e.g.*, depression, anxiety) and cognitive functioning (*e.g.*, processing speed, memory) as a system of interacting symptoms. Given the likely causal role of mental health symptoms on cognitive function in TBI,^10,11,31,32^ we hypothesised that the mental health nodes would act as bridging nodes in mTBI patients. Second, we wanted to simultaneously model brain-behaviour relationships by integrating the behavioural symptom network (comprised of the cognitive functioning and mental health layers) with the brain network. GMVs were selected from regions of the salience and the executive control subnetworks, given their direct involvement in cognitive and mental health deficits across a range of neurological and psychiatric disorders, including TBI.^13–16,33,34^ Our aim was exploratory and consisted primarily in identifying bridging nodes between the mental health, cognitive functioning and GMV networks in mTBI patients.

## METHODS

### Participants and study design

This cohort study used data for 457 mTBI patients drawn from the TRACK-TBI LONG project, a multicentre cohort study conducted across 18 Level 1 trauma centres in the US^28–30,35^. Patients were recruited at each participating institution based on stringent eligibility criteria (as previously described^28–30,35^). Enrolled participants completed a battery of behavioural and self-report outcome assessments and underwent multimodal MRI scanning within each participating institution. For this study, we extracted secondary data from the 6-month follow-up time point. Patients were only included in the final sample if they had *complete* behavioural and neuroimaging data. This study followed the Strengthening the Reporting of Observational Studies in Epidemiology (STROBE) guidelines. See the online supplemental file for full details.

### Mental Health Measures

Patients completed the *BRIEF Symptom Inventory 18 (BSI-18)* and the *Insomnia Severity Index (ISI)*. From these questionnaires, the following measures of symptom severity were selected as nodes for our mental health layer: (1) Anxiety (ANX), (2) Depression (DEP), (3) Somatization (SOM), and Insomnia (INSOM). The Global Severity Index (GSI) score, calculated by summing all ANX, DEP and SOM scores, was also obtained for sample characterization. See the online supplemental file for a full description.

### Cognitive Measures

A core battery of neuropsychological tests from the TRACK-TBI LONG Comprehensive Assessment Battery (https://tracktbi.ucsf.edu/researchers) was also administered. The following measures were selected as nodes for our cognitive layer: (1) Processing speed index (ProcS) (from Symbol Search and Coding subtests of the Wechsler Adult Intelligence Scale IV), (2) psychomotor speed (PsyS) and cognitive flexibility (CogFl) [from the Trail Making Test Part A (TMTA) and Part B (TMTB)], and (3) immediate (vIMM), interference (vINTER), and delayed verbal recall (vDELAY) [from the Rey Auditory Verbal Learning Test II (RAVLT-II)]. See the online supplemental file for a full description.

### Neuroimaging

#### MRI Data Acquisition

We used the T_1_-weighted anatomical MRI data obtained 6 months post-injury. MRI scanning was performed on 3T scanners across all sites. Scanning sequences have been previously described.^30^

#### Grey Matter Volumes

FreeSurfer (6.0)^36^ was used to obtain cortical and subcortical regions of interests (ROI) using the Destrieux atlas.^37,38^ GMV was calculated for each ROI (bilateral average) together with Total Intracranial Volume (TIV). We selected grey-matter ROIs corresponding to the Central Executive (CEN) and Salience (SN) systems.^13–16^ See figure 1 for a visual representation. ROIs from CEN included: (1) Dorsolateral Prefrontal Cortex (dlPFC), (2) Dorsomedial Prefrontal Cortex (dmPFC), (3) Posterior Parietal Cortex (PPC), and (4) Caudate (CAU). ROIs from SN included: (1) Anterior Insula (AntINS), (2) Dorsal Anterior Cingulate Cortex (dACC), (3) Amygdala (AMYG); and (4) Thalamus (THAL). A full list of grey matter parcellations used to create these ROIs is presented in online supplemental table 1.

**Figure 1.**
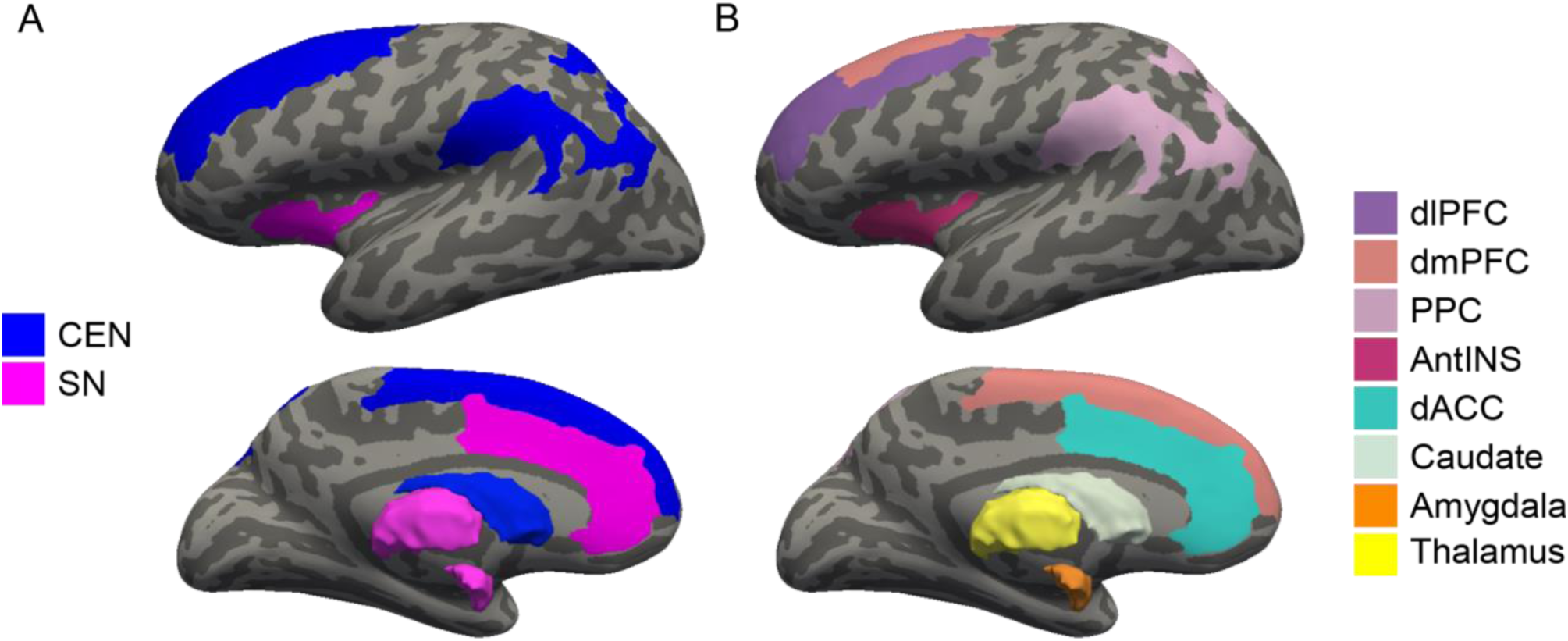
Regions of interest. (A) Visual representation of the Central Executive (CEN) and Salience (SN) networks. (B) Individual cortical and subcortical CEN and SN grey matter regions. Note: dlPFC: Dorsolateral Prefrontal Cortex; dmPFC: Dorsomedial Prefrontal Cortex; PPC: Posterior Parietal Cortex; CAU: Caudate; AntINS: Anterior Insula; dACC: Dorsal Anterior Cingulate Cortex; AMYG: Amygdala; THAL: Thalamus.

### Statistical procedures

#### Network Estimation

Network estimation was conducted in R (4.4.1, “Race for Your Life”; R Core Team, 2024)^39^ using the *bootnet* (1.6),^40^ *igraph* (2.0.3),^41^ and *networktools* (1.5.2)^42^ packages. figure 2 illustrates the workflow to estimate our networks. Nodes included in the networks measured mental health outcomes (4 nodes: ANX, DEP, SOM, ISI), cognitive functioning (6 nodes: ProcS, PsyS, CogFl vIMM, vINTER, vDELAY), and GMV in regions of the CEN and SN (8 nodes – 4 for CEN: dlPFC, dmPFC, PPC, CAU; 4 for SN: AntINS, dACC, AMYG, THAL) (see figure 3 for a list of nodes and labels used throughout). We constructed a bi-layer network with mental health and cognition layers and a tri-layer network with the mental health, cognition and GMV layers. Residualization was used to remove the effect of age from all nodes, and the effect of total intracranial volume (TIV) and site from GMV nodes, prior to network and bridge centrality estimation.^43,44^

**Figure 2.**
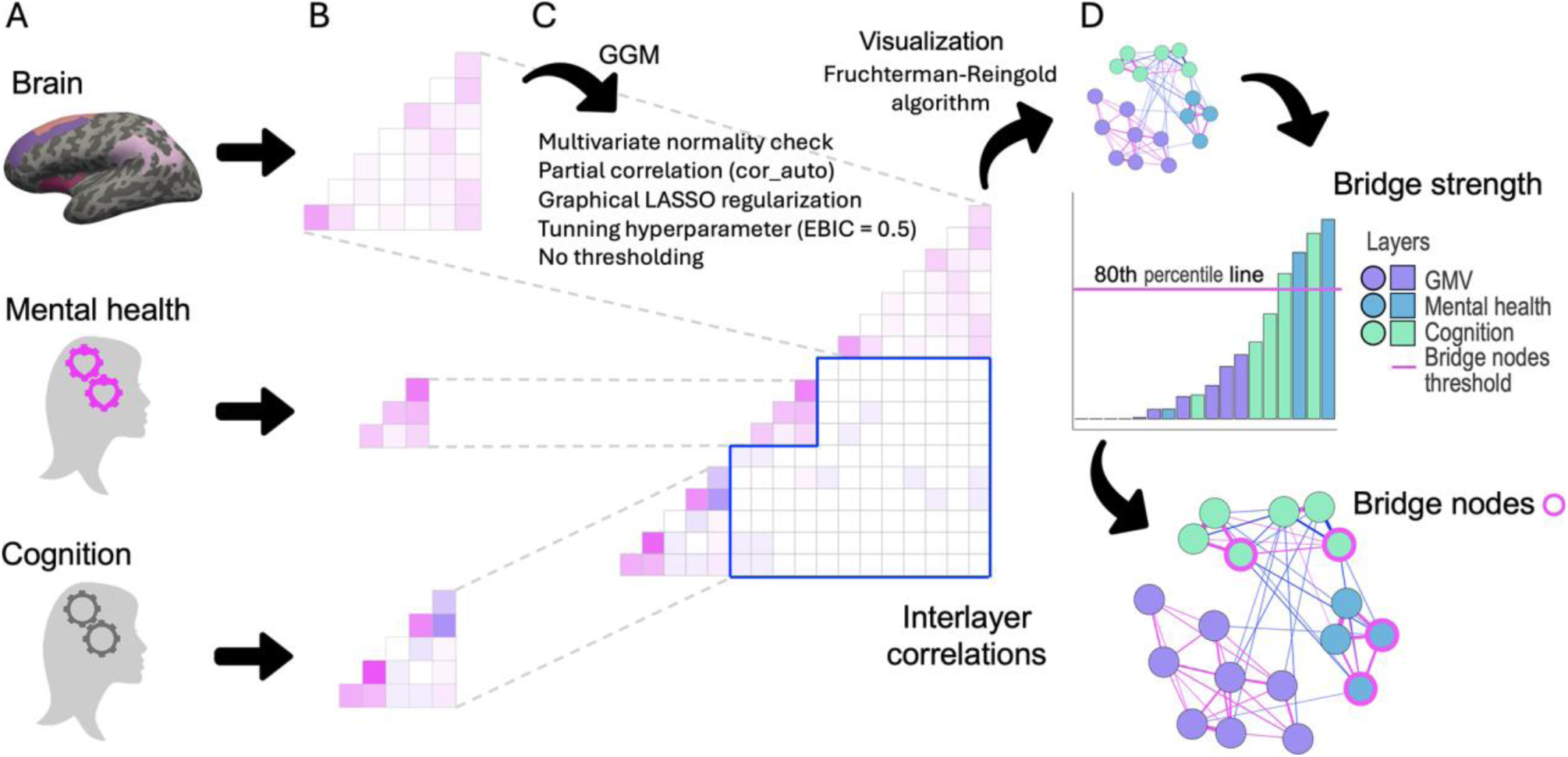
Workflow to estimate multilayer networks and bridge strength. (A) Network layers; (B) Single layer correlations; (C) Multilayer network estimation (GGM, Gaussian Graphical Models); (D) Bridge centrality.

**Figure 3.**
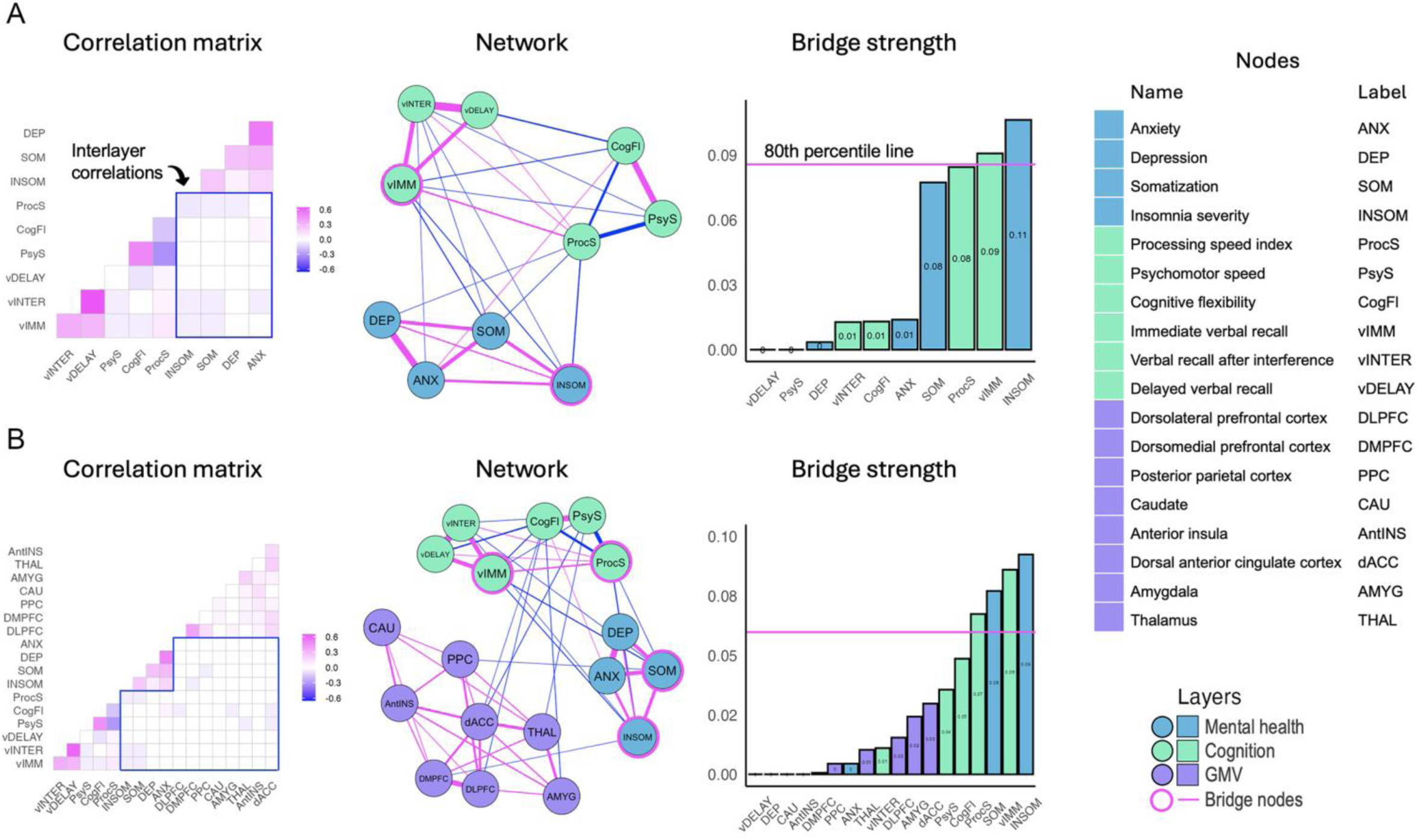
Results of multilayer network analysis in mTBI. (A) Mental health-cognition bi-layer. (B) Mental healthcognition-GMV layer. Note: Weights below 0.05 and −0.05 have been set to ±0.05 in the partial correlation matrices to increase their visibility.

Relationships between nodes (*i.e.*, edges) were estimated as undirected, weighted partial correlations with Gaussian Graphical Models (GGM) using graphical *LASSO* regularisation.^45^ Due to violations of multivariate normality, edge weights for all our networks were estimated using the “cor_auto” function, which automatically computes the correct type of correlation (*e.g.*, Pearson and/or Spearman) for the data. Network density was determined using the Extended Bayesian Information Criterion (EBIC), with the regularisation tuning parameter γ set to 0.5 to control the sparsity of the network. We considered a network density (actual/possible edges) of 50% or more to be high density; 30% to 50% as medium density, and < 30% as low density.^18^ Topological characteristics of each network were visualised in *qgraph* (1.6.5),^46^ with nodes depicted as circles and edges as lines connecting the nodes.^47^ Visualisation of all networks followed the *Fruchterman-Reingold algorithm*.^48^

#### Bridge Strength

*Bridge strength centrality* was estimated to identify nodes that bridge different layers, after assigning nodes to communities defined a priori based on their source layer, *i.e.*, mental health, cognition, or GMV. Bridge strength indicates a node’s total connectivity with layers other than its own. It is computed as the sum of the *absolute value* of every edge weight that connects a node in one community with a node in another community.^24^ Centrality values for each node were then ranked in ascending order. Nodes were designated as bridging nodes by using an 80th percentile threshold on the raw scores of bridge strength.^24^

The stability of bridge strength was assessed by calculating the correlation stability (CS) coefficient between the bridge strength calculated from the initial sample, and estimates calculated after a proportion of participants have been dropped (over 2000 bootstraps). CS coefficients denote the estimated maximum number of individuals that can be dropped to retain, with 95% probability, a correlation of at least 0.70 between the two samples. CS coefficients ≥ 0.50 suggest that the centrality measures are stable and are unlikely to be influenced by variability in sample size.^40^ For CS coefficients between 0.25 and 0.5, centrality measures should be interpreted with caution; if CS falls below 0.25, centrality measures are not stable and should not be interpreted.

## RESULTS

### Demographic and Clinical Characteristics

The final sample included 457 mTBI patients. Table 1 presents all the demographic and clinical characteristics, as well as their scores in the mental health and cognitive measures. None of the total sample reported severe distress (*i.e.*, all GSI scores < 63). A total of 192 mTBI patients reported having experienced either subthreshold (*n* = 112), moderate (*n* = 60) or severe (*n* = 20) symptoms of insomnia.

**Table 1.** Demographic and Clinical Characteristics.

|  | **mTBI (n = 457, 150 females)** |  |  |  |
| --- | --- | --- | --- | --- |
|  | *M* | *SD* | *Min* | *Max* |
| Age | 38.33 | 15.98 | 17 | 83 |
| Days since baseline | 182.11 | 7.98 | 162 | 200 |
| Anxiety (BSI-18) | 2.74 | 4.19 | 0 | 23 |
| Somatization (BSI-18) | 2.54 | 3.71 | 0 | 19 |
| Depression (BSI-18) | 2.83 | 4.37 | 0 | 21 |
| GSI (BSI-18) | 8.11 | 11.02 | 0 | 61 |
| Total Insomnia Severity (ISI) | 7.53 | 6.92 | 0 | 28 |
| Processing speed Index (WAIS-IV PSI) | 102.07 | 15.07 | 53 | 150 |
| Psychomotor speed (TMTA) | 25.70 | 10.85 | 10.47 | 86 |
| Cognitive flexibility (TMTB) | 68.17 | 39.64 | 25 | 330 |
| Immediate verbal recall | 48.09 | 10.67 | 5 | 72 |
| Verbal recall interference | 9.93 | 3.27 | 1 | 15 |
| Delayed verbal recall | 9.70 | 3.49 | 0 | 15 |

*Note:* mTBI: mild Traumatic Brain Injury; BSI-18: Brief Symptom Inventory 18; ISI: Insomnia Severity Index; WAIS-IV PSI: Processing Speed Index of the WAIS-IV; TMTA: Trail Making Test Part A; TMTB: Trail Making Test Part B.

### Network analysis

Descriptive statistics for all multilayer networks in mTBI patients are presented in Table 2 and results of the multilayer network analysis (including the regularised partial correlation matrices) are graphically represented in figure 3.

**Table 2.** Descriptive statistics and stability measures for the Psychometric bi-layer network and the Integrated Brain-Psychometric tri-layer network in mTBI patients.

|  | Edge weight<br>mean (SD), [range] | Non-zero<br>edges/Total No.<br>edges (density) | Bridge<br>Strength<br>stability (CS<sup>†</sup>) |
| --- | --- | --- | --- |
| **Mental health-cognition**<br>**bi-layer network** | 0.06(0.18), [-0.32<br>0.65] | 28 / 45 (62.2%) | 0.27 |
| **Mental health-cognition-**<br>**GMV tri-layer network** | 0.03(0.10), [-0.30<br>0.60] | 53 / 153 (34.6%) | 0.28 |

<sup>†</sup> CS, correlation stability

#### Mental health-cognition bi-layer network of mTBI patients

The density of the mental health-cognition network was high, with 62.2% of possible edges present. This network showed positive and negative partial correlations, with edge weights varying between small and high values (see Table 2). The strongest (positive) edges were between nodes within the same layer, *e.g.*, vINTER-vDELAY (0.65), DEP-ANX (0.53), and PsyS-CogFl (0.49). Interlayer edges were mostly negative, including ProcS-INSOM (−0.07), SOM-vIMM (−0.05), INSOM-vIMM (−0.04), ProcS-SOM (−0.02), vINTER-SOM (−0.01), and ANX-CogFl (0.01). Regarding bridge centrality, two nodes emerged as bridge nodes in the mental health-cognition bi-layer network, with INSOM having the highest bridge strength, followed by vIMM. The stability of these centrality estimates was CS = 0.27.

#### Mental health-cognition-GMV tri-layer network in mTBI patients

The density of the health-cognition-GMV network was medium-to-low, as 34.6% of possible edges were present. It had both positive and negative partial correlations, with edge weights ranging between small and high values (see Table 2). Consistent with the bi-layer network, the strongest edges were within-layer, predominantly positive, and included the edges vINTER-vDELAY (0.60), DEP-ANX (0.50), PsyS-CogFl (0.46), and DLPFC-DMPFC (0.39). In the mental health-cognition-GMV tri-layer network, interlayer the observed edges included ProcS-INSOM (−0.05), vIMM-SOM (−0.05), INSOM-vIMM (−0.04), PsyS-dACC (−0.03), CogFl-AMYGD (−0.02), CogFl-DLPFC (−0.02), vINTER-SOM (−0.01), and PsyS-THAL (−0.01) (a number of additional interlayer edges were observed that had edge weights below an absolute strength of 0.01). Four nodes could be identified as bridge nodes. These included (in descending order): INSOM, vIMM, SOM, and ProcS. Regarding stability, the CS coefficient was 0.28.

## DISCUSSION

In the present study, we constructed for the first time multilayer networks in mTBI patients that integrated self-report questionnaires of mental health, cognitive tests scores from the NIH toolbox, and GMV from anatomical MRI scans. We found four nodes passed the centrality threshold in our multilayer networks, including *insomnia severity*, *immediate verbal recall*, *somatisation*, and *processing speed*. In the next sections, we will discuss these bridging nodes, the clinical implications, limitations of the study and possible avenues for future research.

### Mental health-cognition bi-layer network in mTBI patients

The mental health-cognition network in mTBI patients had a high density. This finding adds to evidence showing that TBI leads to a constellation of interrelated sequelae involving both affective and cognitive symptoms.^10,11,31,32^ This is in agreement with Carmichael and colleagues,^18^ who found that their network of anxiety and depressive symptoms in moderate-to-severe TBI patients had medium density, which confirms the comorbidity and reciprocity of anxiety and depression in this population.^49,50^ Our present findings further show that this is the case not only for anxiety and depression, but other affective symptoms and cognitive deficits as well. This aligns with the view of network psychometrics according to which the effect of a symptom is more likely to spread throughout a dense, strongly interconnected symptom network, potentially leading to a self-sustaining, ingrained mental health disorder.^51^

*Insomnia severity* and *immediate verbal recall* were found to be central nodes that bridge the mental health and cognitive layers. Therefore, our hypothesis that the mental health nodes would act as bridging nodes in the mental health-cognition bi-layer network in mTBI patients was only partially supported in that the node with the highest bridge strength (*insomnia severity*) was a mental health node, but the other bridging node (*immediate verbal recall*) was a cognitive node. Studies have demonstrated that alterations in sleep quality and quantity are commonly found after mTBI.^52,53^ Approximately 50% of mTBI patients report insufficient and disturbed sleep. We similarly found that 42% of our sample experienced either subthreshold, moderate or severe symptoms of insomnia. We also found that *insomnia severity* had strong negative edges with cognitive nodes, including *immediate verbal recall* (the other bridge node in this network) and *processing speed index* (the strongest interlayer edge). Patients with mTBI often experience deficits in *immediate verbal recall* and have a shorter verbal memory span, making it harder to recall a list of words or numbers immediately after hearing them.^54^ Tests used to estimate episodic memory abilities^55^ have revealed high levels of sensitivity and specificity to disorders causing verbal memory dysfunction, including TBI.^56^

Our results align with clinical studies showing that sleep problems post-injury are associated with greater cognitive impairment in mTBI.^57,58^ Future research should continue to examine the relationships between cognitive performance and mental health outcomes after TBI, rather than explore them in isolation. Understanding the interplay between these symptoms is also essential for comprehensive care of TBI patients.^54^ Effective rehabilitation programs addressing verbal memory may result in improvements in mental health.

### Mental health-cognition-GMV tri-layer network in mTBI patients

Bridge centrality analysis revealed the existence of four nodes that served to bridge layers: *insomnia s*everity and *immediate verbal recall (*which were the same as in the mental health-cognitive bi-layer network; see discussion above), as well as *somatization* and *processing speed index.* All four bridge nodes were interconnected, reaffirming their centrality in the multilayer network. Our results highlight the importance of mental health and cognitive functioning in understanding brain-behaviour relationships in mTBI.

Regarding *somatization* (from the mental health layer), previous work indicates that patients with TBI, especially those with mTBI, often report somatic symptoms that are not directly attributable to the injury itself, such as gastrointestinal upset, headaches, dizziness, musculoskeletal pain, and cardiorespiratory complaints.^59^ These symptoms can persist long after the initial injury and can complicate recovery.^60^ *Processing speed index* (from the cognitive layer), which measures perceptual processing speed ability, has been shown to be affected across (complicated) mild, moderate and severe TBI.^61,62^ Slower cognitive processing is well established in mTBI.^6,63^ For example, in their meta-analysis, Frencham and colleagues^63^ found significant deficits in neuropsychological measures, with information-processing speed showing the largest effect.

We observed that *somatization* had strong negative edges with cognitive nodes, including *immediate verbal recall* and *processing speed index* (both of which were bridging nodes), as well as *verbal recall interference*. Studies have demonstrated that the presence of somatic symptoms can exacerbate cognitive deficits, including processing speed.^64^ Conversely, cognitive deficits can increase psychological distress, potentially leading to more pronounced somatic symptoms. Both somatization and processing speed deficits can significantly hinder the recovery process in TBI patients. Understanding and addressing this relationship is crucial for developing effective rehabilitation strategies for TBI patients.

We found no GMV bridge nodes. This result is inconsistent with the multi-layer network study by Simpson-Kent and colleagues,^27^ who showed that bridge brain nodes were stronger than the bridge cognitive nodes in a sample of young struggling learners. This may be due to the low between-layer network density and edge weights in our mTBI sample, particularly between the GMV layer and the other two (mental health and cognition) layers. Future studies should investigate regions beyond those in the CEN and SN networks or use measures other then GMV to assess brain connectivity, like fibre density (from diffusion weighted imaging) or functional connectivity (from functional MRI). However, we did find that the cognitive nodes *processing speed* (from TMTA) and *cognitive flexibility* (from TMTB) had negative edges with *DLPFC*, and *ACC*, both of which have been found to be involved in TMT across lesion-mapping as well as fMRI connectivity studies.^65^ These edges may therefore constitute pathways for the spread of injury effects across brain and behaviour networks in mTBI and warrant further investigation.

### Clinical implications

Bridge centrality measures can inform the development of treatment strategies to improve mental health and cognitive functioning in TBI patients. Specifically, the bridging nodes identified in our cutting-edge multilayer network analyses can be used as targets to develop more customized, efficient, and efficacious treatment programs that target key nodes involved in the mental health symptoms or cognitive deficits of mTBI patients.^23^ For example, a computerized cognitive training program targeting processing speed, such as BrainGames^66^ or Cogmed,^67^ or immediate verbal recall training^68^ may lead to improved cognitive performance, which may result in improvements in mental health in mTBI patients. Treatment can also focus on disturbed sleep as a modifiable treatment target as it has high likelihood of improving outcomes in mTBI patients.^69,70^

### Limitations and conclusions

One limitation of our study is that the findings are based a single time-point. A longitudinal analysis is required to examine changes in the multilayer networks in mTBI patients over time. We did not use other timepoints from the TRACK-TBI dataset (1 week and 12 months post-injury) due to the high number of missing cells for the behavioural and/or brain data of the other time points. Advances in statistical modelling will enable us to conduct longitudinal multilayer network analyses in future work, which will allow inferences about the time-related dynamics of these multilayer networks. Another limitation is that our study included mTBI patients only. Future multilayer network studies should include samples at different severity levels of TBI. Also important will be to use data from a broad spectrum of TBI populations, such as those from the ENIGMA TBI working group.^71–73^

Another shortcoming of our study is that, while both the bi- and tri-layer networks had sufficient stability, they should be interpreted with caution as their CS was below 0.5. This may stem from the high degree of heterogeneity that characterizes mTBI patients. While the sample size in the present multilayer network study is considerably larger than those commonly used in standard univariate neuroimaging studies in TBI patients,^74–76^ our stability estimates suggest that even larger samples be used for this type of analysis so that node centrality estimates can be generated that are stable.

Despite these limitations, to the best of our knowledge, ours is the first study to use a multilayer network approach that simultaneously models brain-behaviour relationships in mTBI patients. Using bridge strength centrality, we were able to identify nodes, including *insomnia severity, immediate verbal memory*, *somatization*, and *processing speed,* that work as bridges across network layers. These bridges can be considered as potential training targets and can help pinpoint strategies (*e.g.*, using computerized cognitive training programs, sleep-promoting interventions) to improve outcomes in mTBI patients.

## Contributors

J.F.D.D, M.S and K.C. contributed to the conception and design of the study. J.F.D.D. and M.S. contributed to the analysis of data. J.F.D.D, M.S, L.F., J.G, I.L.S., P.I., A.I., K.C contributed to drafting the manuscript. All authors approved the manuscript.

## Funders

KC is supported by a Veski Fellowship. The Victorian Near-miss Award Pilot is administered by Veski for the Victorian Health and Medical Research Workforce Project on behalf of the Victorian Government and the Association of Australian Medical Research Institutes.

## Competing Interests

The authors report no competing interests.

## Patient consent for publication

Written consent was granted by all enrolled participants and/or their legally authorised representatives prior to participation in the study.

## Ethics approval

Ethics for recruitment and assessment were approved by the University of California, San Francisco, and Institutional Review Boards for each participating site.

## Data Availability Statement

Data used for this manuscript are available upon reasonable request. Requests must be submitted as proposals to the study leadership of TRACK-TBI.

## Notes

### Competing Interest Statement

The authors have declared no competing interest.

### Author Declarations

This study was approved by the Institutional Review Boards of Baylor College of Medicine; Massachusetts General Hospital/Spaulding Rehabilitation Hospital; University of California, San Francisco; University of Cincinnati; University of Maryland; University of Miami; University of Pittsburgh; University of Texas, Austin; University of Texas, Southwestern; University of Washington; Virginia Commonwealth University; University of Pennsylvania; Emory University; Medical College of Wisconsin; University of Utah; Indiana University; Hennepin County Medical Center; and Denver Health Medical Center - Craig Hospital.

## REFERENCES

1. Wu L, Li Y, Sun M, Ye P, Zhang Z, Liu W. Global, regional, and national burdens of mild traumatic brain injuries from 1990-2019: findings from the Global Burden of Disease Study 2019 - a cross-sectional study. Int J Surg. Jun 24 2024;doi:10.1097/js9.0000000000001837

2. Carroll EL, Outtrim JG, Forsyth F, et al. Mild traumatic brain injury recovery: a growth curve modelling analysis over 2 years. J Neurol. Nov 2020;267(11):3223–3234. doi:10.1007/s00415-020-09979-x

3. Miotto EC, Cinalli FZ, Serrao VT, Benute GG, Lucia MC, Scaff M. Cognitive deficits in patients with mild to moderate traumatic brain injury. Arq Neuropsiquiatr. Dec 2010;68(6):862–8. doi:10.1590/s0004-282x2010000600006

4. Barman A, Chatterjee A, Bhide R. Cognitive Impairment and Rehabilitation Strategies After Traumatic Brain Injury. Indian J Psychol Med. May-Jun 2016;38(3):172–81. doi:10.4103/0253-7176.183086

5. Tsai YC, Liu CJ, Huang HC, et al. A Meta-analysis of Dynamic Prevalence of Cognitive Deficits in the Acute, Subacute, and Chronic Phases After Traumatic Brain Injury. J Neurosci Nurs. Apr 1 2021;53(2):63–68. doi:10.1097/jnn.0000000000000570

6. Hacker D, Jones CA, Yasin E, et al. Cognitive Outcome After Complicated Mild Traumatic Brain Injury: A Literature Review and Meta-Analysis. J Neurotrauma. Oct 2023;40(19-20):1995–2014. doi:10.1089/neu.2023.0020

7. Imms P, Chowdhury NF, Chaudhari NN, et al. Prediction of cognitive outcome after mild traumatic brain injury from acute measures of communication within brain networks. Cortex. Feb 2024;171:397–412. doi:10.1016/j.cortex.2023.10.022

8. Howlett JR, Nelson LD, Stein MB. Mental Health Consequences of Traumatic Brain Injury. Biol Psychiatry. Mar 1 2022;91(5):413–420. doi:10.1016/j.biopsych.2021.09.024

9. Karakurt G, Whiting K, Jones SE, Lowe MJ, Rao SM. Brain Injury and Mental Health Among the Victims of Intimate Partner Violence: A Case-Series Exploratory Study. Front Psychol. 2021;12:710602. doi:10.3389/fpsyg.2021.710602

10. Keatley ES, Bombardier CH, Watson E, et al. Cognitive Performance, Depression, and Anxiety 1 Year After Traumatic Brain Injury. J Head Trauma Rehabil. May-Jun 01 2023;38(3):E195–e202. doi:10.1097/htr.0000000000000819

11. Uiterwijk D, Stargatt R, Humphrey S, Crowe SF. The Relationship Between Cognitive Functioning and Symptoms of Depression, Anxiety, and Post-Traumatic Stress Disorder in Adults with a Traumatic Brain Injury: a Meta-Analysis. Neuropsychol Rev. Dec 2022;32(4):758–806. doi:10.1007/s11065-021-09524-1

12. Amgalan A, Maher AS, Imms P, Ha MY, Fanelle TA, Irimia A. Functional Connectome Dynamics After Mild Traumatic Brain Injury According to Age and Sex. Front Aging Neurosci. 2022;14:852990. doi:10.3389/fnagi.2022.852990

13. Li X, Jia X, Liu Y, et al. Brain dynamics in triple-network interactions and its relation to multiple cognitive impairments in mild traumatic brain injury. Cereb Cortex. May 24 2023;33(11):6620–6632. doi:10.1093/cercor/bhac529

14. Liu H, Zhang G, Zheng H, et al. Dynamic Dysregulation of the Triple Network of the Brain in Mild Traumatic Brain Injury and Its Relationship With Cognitive Performance. J Neurotrauma. Apr 2024;41(7-8):879–886. doi:10.1089/neu.2022.0257

15. Menon V. Large-scale brain networks and psychopathology: a unifying triple network model. Trends Cogn Sci. Oct 2011;15(10):483–506. doi:10.1016/j.tics.2011.08.003

16. van der Horn HJ, Dodd AB, Wick TV, et al. Neural correlates of cognitive control deficits in pediatric mild traumatic brain injury. Hum Brain Mapp. Dec 1 2023;44(17):6173–6184. doi:10.1002/hbm.26504

17. Borsboom D, Fried EI, Epskamp S, et al. False alarm? A comprehensive reanalysis of “Evidence that psychopathology symptom networks have limited replicability” by Forbes, Wright, Markon, and Krueger (2017). 2017.

18. Carmichael J, Hicks AJ, Gould KR, Spitz G, Ponsford J. Network analysis of anxiety and depressive symptoms one year after traumatic brain injury. Psychiatry Res. Aug 2023;326:115310. doi:10.1016/j.psychres.2023.115310

19. Schubert A-L, Frischkorn GT. Neurocognitive psychometrics of intelligence: How measurement advancements unveiled the role of mental speed in intelligence differences. Current Directions in Psychological Science. 2020;29(2):140–146.

20. van der Maas H, Savi AO, Hofman A, Kan KJ, Marsman M. The network approach to general intelligence. 2021.

21. Yang T, Guo Z, Cao X, et al. Network analysis of anxiety and depression in the functionally impaired elderly. Front Public Health. 2022;10:1067646. doi:10.3389/fpubh.2022.1067646

22. Tiego J, Fornito A. Putting behaviour back into brain–behaviour correlation analyses. Aperture Neuro. 2022:1–4.

23. Blanken TF, Bathelt J, Deserno MK, Voge L, Borsboom D, Douw L. Connecting brain and behaviour in clinical neuroscience: A network approach. Neurosci Biobehav Rev. Nov 2021;130:81–90. doi:10.1016/j.neubiorev.2021.07.027

24. Jones PJ, Ma R, McNally RJ. Bridge Centrality: A Network Approach to Understanding Comorbidity. Multivariate Behav Res. Mar-Apr 2021;56(2):353–367. doi:10.1080/00273171.2019.1614898

25. Hilland E, Landrø NI, Kraft B, et al. Exploring the links between specific depression symptoms and brain structure: A network study. Psychiatry Clin Neurosci. Mar 2020;74(3):220–221. doi:10.1111/pcn.12969

26. Bathelt J, Geurts HM, Borsboom D. More than the sum of its parts: Merging network psychometrics and network neuroscience with application in autism. Netw Neurosci. Jun 2022;6(2):445–466. doi:10.1162/netn_a_00222

27. Simpson-Kent IL, Fried EI, Akarca D, et al. Bridging Brain and Cognition: A Multilayer Network Analysis of Brain Structural Covariance and General Intelligence in a Developmental Sample of Struggling Learners. J Intell. Jun 15 2021;9(2)doi:10.3390/jintelligence9020032

28. Bryant AM, Rose NB, Temkin NR, et al. Profiles of Cognitive Functioning at 6 Months After Traumatic Brain Injury Among Patients in Level I Trauma Centers: A TRACK-TBI Study. JAMA Netw Open. Dec 1 2023;6(12):e2349118. doi:10.1001/jamanetworkopen.2023.49118

29. Palacios EM, Yuh EL, Mac Donald CL, et al. Diffusion Tensor Imaging Reveals Elevated Diffusivity of White Matter Microstructure that Is Independently Associated with Long-Term Outcome after Mild Traumatic Brain Injury: A TRACK-TBI Study. J Neurotrauma. Oct 2022;39(19-20):1318–1328. doi:10.1089/neu.2021.0408

30. Sibilia F, Custer RM, Irimia A, Sepehrband F, Toga AW, Cabeen RP. Life After Mild Traumatic Brain Injury: Widespread Structural Brain Changes Associated With Psychological Distress Revealed With Multimodal Magnetic Resonance Imaging. Biol Psychiatry Glob Open Sci. Jul 2022;3(3):374–385. doi:10.1016/j.bpsgos.2022.03.004

31. Silver JM, McAllister TW, Arciniegas DB. Depression and cognitive complaints following mild traumatic brain injury. Am J Psychiatry. Jun 2009;166(6):653–61. doi:10.1176/appi.ajp.2009.08111676

32. Silverberg ND, Iverson GL. Etiology of the post-concussion syndrome: Physiogenesis and Psychogenesis revisited. NeuroRehabilitation. 2011;29(4):317–29. doi:10.3233/nre-2011-0708

33. Menon B. Towards a new model of understanding - The triple network, psychopathology and the structure of the mind. Med Hypotheses. Dec 2019;133:109385. doi:10.1016/j.mehy.2019.109385

34. Menon V, D’Esposito M. The role of PFC networks in cognitive control and executive function. Neuropsychopharmacology. Jan 2022;47(1):90–103. doi:10.1038/s41386-021-01152-w

35. Yue JK, Vassar MJ, Lingsma HF, et al. Transforming Research and Clinical Knowledge in Traumatic Brain Injury Pilot: Multicenter Implementation of the Common Data Elements for Traumatic Brain Injury. Journal of Neurotrauma. 2013/11/15 2013;30(22):1831–1844. doi:10.1089/neu.2013.2970

36. Fischl B. FreeSurfer. Neuroimage. Aug 15 2012;62(2):774–81. doi:10.1016/j.neuroimage.2012.01.021

37. Destrieux C, Fischl B, Dale A, Halgren E. Automatic parcellation of human cortical gyri and sulci using standard anatomical nomenclature. Neuroimage. Oct 15 2010;53(1):1–15. doi:10.1016/j.neuroimage.2010.06.010

38. Fischl B, van der Kouwe A, Destrieux C, et al. Automatically parcellating the human cerebral cortex. Cereb Cortex. Jan 2004;14(1):11–22. doi:10.1093/cercor/bhg087

39. R Core Team. Package ‘parallel’. 2024.

40. Epskamp S, Borsboom D, Fried EI. Estimating psychological networks and their accuracy: A tutorial paper. Behav Res Methods. Feb 2018;50(1):195–212. doi:10.3758/s13428-017-0862-1

41. Csardi G, Nepusz T. The igraph software. Complex syst. 2006;1695:1–9.

42. Jones P. Networktools: Tools for Identifying Important Nodes in Networks. *R package version 152.* 2024;1doi:https://CRAN.R-project.org/package=networktools

43. Bethlehem RAI, Seidlitz J, White SR, et al. Brain charts for the human lifespan. Nature. Apr 2022;604(7906):525–533. doi:10.1038/s41586-022-04554-y

44. Storsve AB, Fjell AM, Tamnes CK, et al. Differential longitudinal changes in cortical thickness, surface area and volume across the adult life span: regions of accelerating and decelerating change. J Neurosci. Jun 18 2014;34(25):8488–98. doi:10.1523/jneurosci.0391-14.2014

45. Friedman J, Hastie T, Tibshirani R. Sparse inverse covariance estimation with the graphical lasso. Biostatistics. Jul 2008;9(3):432–41. doi:10.1093/biostatistics/kxm045

46. Epskamp S, Cramer AOJ, Waldorp LJ, Schmittmann VD, Borsboom D. qgraph: Network Visualizations of Relationships in Psychometric Data. Journal of Statistical Software. 05/24 2012;48(4):1–18. doi:10.18637/jss.v048.i04

47. Epskamp S, Waldorp LJ, Mõttus R, Borsboom D. The Gaussian Graphical Model in Cross-Sectional and Time-Series Data. Multivariate Behav Res. Jul-Aug 2018;53(4):453–480. doi:10.1080/00273171.2018.1454823

48. Fruchterman TM, Reingold EM. Graph drawing by force-directed placement. Software: Practice and experience. 1991;21(11):1129–1164.

49. Gould KR, Ponsford JL, Johnston L, Schönberger M. The nature, frequency and course of psychiatric disorders in the first year after traumatic brain injury: a prospective study. Psychol Med. Oct 2011;41(10):2099–109. doi:10.1017/s003329171100033x

50. Wang B, Zeldovich M, Rauen K, et al. Longitudinal Analyses of the Reciprocity of Depression and Anxiety after Traumatic Brain Injury and Its Clinical Implications. J Clin Med. Nov 28 2021;10(23)doi:10.3390/jcm10235597

51. Borsboom D. A network theory of mental disorders. World psychiatry. 2017;16(1):5–13.

52. Montgomery MC, Baylan S, Gardani M. Prevalence of insomnia and insomnia symptoms following mild-traumatic brain injury: A systematic review and meta-analysis. Sleep Med Rev. Feb 2022;61:101563. doi:10.1016/j.smrv.2021.101563

53. Wickwire EM, Schnyer DM, Germain A, et al. Sleep, Sleep Disorders, and Circadian Health following Mild Traumatic Brain Injury in Adults: Review and Research Agenda. J Neurotrauma. Nov 15 2018;35(22):2615–2631. doi:10.1089/neu.2017.5243

54. Vakil E, Greenstein Y, Weiss I, Shtein S. The Effects of Moderate-to-Severe Traumatic Brain Injury on Episodic Memory: a Meta-Analysis. Neuropsychol Rev. Sep 2019;29(3):270–287. doi:10.1007/s11065-019-09413-8

55. Rabin LA, Barr WB, Burton LA. Assessment practices of clinical neuropsychologists in the United States and Canada: a survey of INS, NAN, and APA Division 40 members. Arch Clin Neuropsychol. Jan 2005;20(1):33–65. doi:10.1016/j.acn.2004.02.005

56. Kennedy E, Liebel SW, Lindsey HM, et al. Verbal Learning and Memory Deficits across Neurological and Neuropsychiatric Disorders: Insights from an ENIGMA Mega Analysis. Brain Sci. Jun 29 2024;14(7) doi:10.3390/brainsci14070669

57. Kalmbach DA, Conroy DA, Falk H, et al. Poor sleep is linked to impeded recovery from traumatic brain injury. Sleep. Oct 1 2018;41(10)doi:10.1093/sleep/zsy147

58. Werner JK, Shahim P, Pucci JU, et al. Poor sleep correlates with biomarkers of neurodegeneration in mild traumatic brain injury patients: a CENC study. Sleep. Jun 11 2021;44(6)doi:10.1093/sleep/zsaa272

59. Stenberg J, Karr JE, Terry DP, et al. Change in self-reported cognitive symptoms after mild traumatic brain injury is associated with changes in emotional and somatic symptoms and not changes in cognitive performance. Neuropsychology. 2020;34(5):560.

60. Stubbs JL, Green KE, Silverberg ND, et al. Atypical Somatic Symptoms in Adults With Prolonged Recovery From Mild Traumatic Brain Injury. Front Neurol. 2020;11:43. doi:10.3389/fneur.2020.00043

61. Carlozzi NE, Kirsch NL, Kisala PA, Tulsky DS. An examination of the Wechsler Adult Intelligence Scales, Fourth Edition (WAIS-IV) in individuals with complicated mild, moderate and Severe traumatic brain injury (TBI). Clin Neuropsychol. 2015;29(1):21–37. doi:10.1080/13854046.2015.1005677

62. Donders J, Strong CA. Clinical utility of the Wechsler Adult Intelligence Scale-Fourth Edition after traumatic brain injury. Assessment. Feb 2015;22(1):17–22. doi:10.1177/1073191114530776

63. Frencham KA, Fox AM, Maybery MT. Neuropsychological studies of mild traumatic brain injury: a meta-analytic review of research since 1995. J Clin Exp Neuropsychol. Apr 2005;27(3):334–51. doi:10.1080/13803390490520328

64. Calvillo M, Irimia A. Neuroimaging and Psychometric Assessment of Mild Cognitive Impairment After Traumatic Brain Injury. Front Psychol. 2020;11:1423. doi:10.3389/fpsyg.2020.01423

65. Varjacic A, Mantini D, Demeyere N, Gillebert CR. Neural signatures of Trail Making Test performance: Evidence from lesion-mapping and neuroimaging studies. Neuropsychologia. Jul 1 2018;115:78–87. doi:10.1016/j.neuropsychologia.2018.03.031

66. Verhelst H, Giraldo D, Vander Linden C, Vingerhoets G, Jeurissen B, Caeyenberghs K. Cognitive Training in Young Patients With Traumatic Brain Injury: A Fixel-Based Analysis. Neurorehabil Neural Repair. Oct 2019;33(10):813–824. doi:10.1177/1545968319868720

67. Caeyenberghs K, Metzler-Baddeley C, Foley S, Jones DK. Dynamics of the Human Structural Connectome Underlying Working Memory Training. J Neurosci. Apr 6 2016;36(14):4056–66. doi:10.1523/jneurosci.1973-15.2016

68. Lambez B, Vakil E. The effectiveness of memory remediation strategies after traumatic brain injury: Systematic review and meta-analysis. Ann Phys Rehabil Med. Sep 2021;64(5):101530. doi:10.1016/j.rehab.2021.101530

69. Killgore WDS, Vanuk JR, Shane BR, Weber M, Bajaj S. A randomized, double-blind, placebo-controlled trial of blue wavelength light exposure on sleep and recovery of brain structure, function, and cognition following mild traumatic brain injury. Neurobiol Dis. Feb 2020;134:104679. doi:10.1016/j.nbd.2019.104679

70. Ouellet MC, Morin CM. Efficacy of cognitive-behavioural therapy for insomnia associated with traumatic brain injury: a single-case experimental design. Arch Phys Med Rehabil. Dec 2007;88(12):1581–92. doi:10.1016/j.apmr.2007.09.006

71. Dennis EL, Caeyenberghs K, Hoskinson KR, et al. White matter disruption in pediatric traumatic brain injury: Results from ENIGMA pediatric moderate to severe traumatic brain injury. Neurology. 2021;97(3):e298–e309.

72. Keleher F, Lindsey HM, Kerestes R, et al. Multimodal Analysis of Secondary Cerebellar Alterations After Pediatric Traumatic Brain Injury. JAMA Netw Open. Nov 1 2023;6(11):e2343410. doi:10.1001/jamanetworkopen.2023.43410

73. Olsen A, Babikian T, Bigler ED, et al. Toward a global and reproducible science for brain imaging in neurotrauma: the ENIGMA adult moderate/severe traumatic brain injury working group. Brain Imaging Behav. Apr 2021;15(2):526–554. doi:10.1007/s11682-020-00313-7

74. Livny, A., Biegon, A., Kushnir, T., Harnof, S., Hoffmann, C., Fruchter, E., & Weiser, M. (2017). Cognitive deficits post-traumatic brain injury and their association with injury severity and gray matter volumes. Journal of Neurotrauma, 34(7), 1466–1472.

75. Hellstrøm T, Westlye LT, Kaufmann T, et al. White matter microstructure is associated with functional, cognitive and emotional symptoms 12 months after mild traumatic brain injury. Scientific reports. 2017;7(1):13795.

76. Rostowsky KA, Irimia A. Acute cognitive impairment after traumatic brain injury predicts the occurrence of brain atrophy patterns similar to those observed in Alzheimer’s disease. Geroscience. Aug 2021;43(4):2015–2039. doi:10.1007/s11357-021-00355-9

