## supplemental for "Bridging mental health, cognition and the brain in mild traumatic brain injury: A multilayer network analysis of the TRACK-TBI study"

### Methods

#### *Study Design and Procedure*

For this cohort study, eligible mild traumatic brain injury (mTBI) patients were identified if they were: (1) aged 17 years and older, (2) presented with a documented case of TBI within 24 hours after injury, (3) obtained a clinical computed tomography (CT) scan within 24 hours after injury; (4) exhibited a disorder of consciousness, and (5) reported a total Glasgow Coma Scale (GCS) score of 13-15 points (indicating presence of mild TBI) upon admission. Participants were excluded if they were incarcerated, pregnant, or diagnosed with any of the following: (1) penetrating TBI, (2) psychiatric disorders, (3) neurological disorders; and (3) non-English-speaking. Participants were enrolled between March 2, 2014, and July 27, 2018. Data were analysed between January and August 2024.

#### *Mental Health Measures*

*BRIEF Symptom Inventory 18 (BSI-18)*

Mental-health symptoms were assessed using the Brief Symptom Inventory 18 (BSI-18),[1] an 18-item self-report questionnaire assessing the severity of psychiatric distress symptoms across three subdomains: (1) Anxiety (*ANX*; distress relating to symptoms of anxiety), (2) Depression (*DEP*; distress relating to symptoms of depression), and (3) Somatization (*SOM*; distress relating to bodily dysfunction, e.g., nausea, faintness). Total subdomain scores ranged between 0-24, with higher scores indicating greater severity of distress for that subdomain. Total ANX, DEP, and SOM scores for each participant were included as nodes for our affective layer (see Table S2 for a full list of all nodes and labels as well as their source). The BSI-18 also enables the quantification of a Global Severity Index (GSI) score, which was obtained by summing all subdomain scores. The GSI total score ranges between 0-72, with scores $\geq 63$ indicative of having significant distress.[1] This GSI score was used for characterization of our sample.

*Insomnia Severity Index (ISI)*

The severity of insomnia symptoms was assessed using the Insomnia Severity Index (ISI), a brief screening tool composed of seven 4-point Likert scale items evaluating subjective perception of insomnia symptoms over the past two weeks.[2] A total score of insomnia severity (*INSOM*) was calculated from the sum of all seven items, with scores ranging from 0-28. Subthreshold insomnia is indicated by scores between 8-14, whilst moderate and severe clinical insomnia is given as scores between 15-21 and 22-28, respectively.[2] *INSOM* was included as a node in our affective layer.

***Cognitive Measures***

*WAIS-IV Processing Speed Index*

The Symbol Search and Coding subtests of the Wechsler Adult Intelligence Scale, Fourth Edition (WAIS-IV) are measures of perceptual processing speed (comprising the WAIS-IV Processing speed Index[3]). During the *Symbol Search*, participants were presented with a series of two visual symbols to the left of a set of five other symbols to which they were instructed to use a pencil to mark 'yes' in the checkbox if the five symbols contained either of the two target symbols, and 'no' if there was no match. Each item was presented sequentially as a row and participants completed as many items as they could within two minutes. An overall accuracy score was calculated from the sum of the total correct positive and negative responses. In the *Coding* subtest, participants were presented with a series of boxes numbered from 1-9 with each number corresponding to a unique symbol. Participants were then instructed to draw the symbol that matches each number in another set of empty numbered boxes (1-9). An overall raw score was obtained from the number of correct responses after a maximum duration of two minutes. Lastly, a total Processing Speed Index (*ProcS*) composite score was calculated by summing the scale scores from both subtests. Higher scores denote better processing speed ability. *ProcS* was included as a node in our psychometric network.

*Trail Making Test Parts A & B*

The Trail Making Test Parts A and B are commonly used neuropsychological measures of psychomotor speed and mental flexibility.[4,5] In *Part A*, participants were instructed to connect a series of 25 numbered circles in ascending order as quickly as possible with limited feedback by the examiner on any errors made. Participants exceeding 100 seconds were discontinued from the task. The time taken to complete the task was calculated as a reaction time (RT) to assess overall psychomotor speed (*PsyS*), with lower scores indicative of faster psychomotor speed. In *Part B*, two types of stimuli were presented; (1) a set of circles numbered from 1-13 and (2) a set of circles with letters from A-L. Participants were again tasked to link each circle in an ascending pattern, but with the added requirement of alternating between the different stimulus patterns (i.e., switching between letters and numbers), thus measuring cognitive flexibility (*CogFl*). Participants exceeding 300 seconds were discontinued. Same as in Part A, performance in Part B was calculated as an RT measure. Both *PsyS* and *CogFl* were included as nodes in our cognitive network.

*Rey Auditory and Verbal Learning Test II*

The Rey Auditory Verbal Learning Test II (RAVLT-II) evaluates verbal learning and episodic memory.[6] The test included three parts. In the first part, participants were asked to recall a list of 15 unrelated words (List A) repeated across five different trials (Trials 1-5). A total measure of immediate verbal recall (*vIMM*) was obtained from summing together the number of words correctly recalled for each trial. In the next part (Trial 6), participants were given an interference set of 15 new words (List B), after which they were asked to recall the original words from List A without mentioning any words belonging to the interference set.

Performance during Trial 6 was calculated as the number of correctly recalled List A words after the administration of the interference set (*vINTER*). In the final part of the test (Trial 7), participants were again asked to recall as many words as they can remember from List A. However, this time the trial was conducted after a 20-minute delay (during which other assessments were administered). The number of correct List A words was then recorded to assess the effect of delay on episodic verbal recall (*vDELAY*). *vIMM*, *vINTER* and *vDELAY* were used as nodes in our psychometric network.

**Tables**

**Supplemental table 1**. List of Destrieux gyral and sulcal parcellations used to generate the ROIs in our Integrated Brain-Behavioral network.

| **Destrieux label** | **ROI** | **Network** |
|---|---|---|
| Middle frontal gyrus | dlPFC | CEN |
| Middle frontal sulcus | dlPFC | CEN |
| Superior frontal gyrus | dmPFC | CEN |
| Superior frontal sulcus | dmPFC | CEN |
| Superior parietal lobule | PPC | CEN |
| Angular gyrus | PPC | CEN |
| Supramarginal gyrus | PPC | CEN |
| Caudate | CAU | CEN |
| Short insular gyri | AntINS | SN |
| Anterior segment circular sulcus of the insula | AntINS | SN |
| Anterior cingulate gyrus and sulcus | dACC | SN |
| Middle-anterior cingulate gyrus and sulcus | dACC | SN |
| Amygdala | AMYG | SN |
| Thalamus | THAL | SN |

**Supplemental table 2**. List of nodes included in the networks.

| Layer | Node Name | Node Label | Source |
|---|---|---|---|
| Mental health | Anxiety | ANX | BSI-18 |
| Mental health | Depression | DEP | BSI-18 |
| Mental health | Somatization | SOM | BSI-18 |
| Mental health | Insomnia severity | INSOM | ISI |
| Cognition | Processing speed index | ProcS | WAIS-IV |
| Cognition | Psychomotor speed | PsyS | TMTA |
| Cognition | Cognitive flexibility | CogFl | TMTB |
| Cognition | Immediate verbal recall | vIMM | RAVLT-II |
| Cognition | Verbal recall after interference | vINTER | RAVLT-II |
| Cognition | Delayed verbal recall | vDELAY | RAVLT-II |
| GMV | Dorsolateral prefrontal cortex | DLPFC | FS volume |
| GMV | Dorsomedial prefrontal cortex | DMPFC | FS volume |
| GMV | Posterior parietal cortex | PPC | FS volume |
| GMV | Caudate | CAU | FS volume |
| GMV | Anterior insula | AntINS | FS volume |
| GMV | Dorsal anterior cingulate cortex | dACC | FS volume |
| GMV | Amygdala | AMYG | FS volume |
| GMV | Thalamus | THAL | FS volume |

BSI-18, The Brief Symptom Inventory 18; ISI, Insomnia Severity Index; WAIS-IV, Wechsler Adult Intelligence Scale, Fourth Edition; TMT, Trail Making Test Parts A and B; RAVLT-II , Rey Auditory Verbal Learning Test II; FS, FreeSurfer.

**References**

1. Derogatis, L. R., & Melisaratos, N. (1983). The Brief Symptom Inventory: An introductory report. *Psychological Medicine, 13*(3), 595–605. https://doi.org/10.1017/S0033291700048017
2. Morin, C. M., Belleville, G., Bélanger, L., & Ivers, H. (2011). The Insomnia Severity Index: psychometric indicators to detect insomnia cases and evaluate treatment response. Sleep, 34(5), 601–608. https://doi.org/10.1093/sleep/34.5.601
3. Wechsler, D. (2008). *Wechsler Adult Intelligence Scale--Fourth Edition (WAIS-IV)* https://doi.org/10.1037/t15169-000
4. Bowie, C. R., & Harvey, P. D. (2006). Administration and interpretation of the Trail Making Test. *Nature protocols*, *1*(5), 2277–2281. https://doi.org/10.1038/nprot.2006.390
5. Salthouse T. A. (2011). What cognitive abilities are involved in trail-making performance? *Intelligence*, *39*(4), 222–232. https://doi.org/10.1016/j.intell.2011.03.001
6. Schmidt M (1996). Rey auditory and verbal learning test: A handbook. Western Psychological Services
